# Prompt-engineering improves clinical safety of large language models for opioid equipotency conversion

**DOI:** 10.64898/2026.05.06.26352590

**Authors:** Tanya M. Marton, David Corpman, Lytia Lai, Rodney A. Gabriel, Yian Chen

## Abstract

**Background:** Large language models (LLMs) are increasingly used in medical education and clinical decision-making, but their reliability in high-risk medication dosing remains unclear. Opioid rotation is a common task requiring precise calculations where errors may result in overdose or inadequate pain relief.

**Methods:** Thirteen LLMs were tested using an API-based framework to ensure independent queries across trials. First, fictional clinical scenarios were tested to simulate real-world clinical situations involving opioid rotation; to test the effects of changes in wording, scenarios were revised into 4 “vignettes” showing the same clinical situation. Next, opioid pairs were tested with a random-dose paradigm across a clinically-pertinent range (5-120 mg daily morphine equivalents). LLM outputs were compared with expected values derived from reference standards. Accuracy was assessed using predefined safety thresholds: tight accuracy (0.85–1.15x expected dose) and broad accuracy (0.6–1.7x). We tested models naively and with prompts augmented with reference tables and unit explanations.

**Results:** Naive models generally exhibited low tight-range accuracy across opioid pairs. For any given opioid pair, each model would consistently produce similar incorrect conversion ratios despite wide variability across opioid pairs and language models. Vignette wording changes accounted for 76% of within-scenario response variance. Reference-based prompt augmentation significantly improved performance, with over half of models achieving high proportions of conversions within tight accuracy for morphine-equivalent conversions.

**Conclusions:** While commercial LLMs demonstrated variable accuracy in the native state, prompt augmentation significantly improved their performance.

## Introduction

While opioid use in the United States has decreased over the past decade ^1^, particularly with efforts to limit prescriptions, they remain widely-used with over 125 million prescriptions dispensed in 2023 ^2^. Opioid rotation remains an important cornerstone in stewardship, permitting possible reduction of side effects or improving effectiveness in the context of tolerance ^3^. However, safe opioid rotation requires an accurate understanding of morphine equivalency and careful measurements of equipotency between different agents. Opioid equivalence has been calculated through a variety of means, including online conversion calculators and equivalence tables ^4^, but studies have shown substantial variation and thus potential for patient harm. Providers are also apt to arrive at different conclusions regarding equivalency. Rennick et al. electronically surveyed physicians, nurses and other advanced providers asking about estimated morphine equivalents for a variety of synthetic opioids; large standard deviations were reported for each conversion ^5^. Accuracy of opioid conversion is essential in reducing risk of overdose and minimizing the harmful effects of withdrawal. Opioid rotation or conversion has been a problematic topic for trainees and even practicing physicians ^6–8^. Several case reports have illustrated dramatic errors due to a lack of understanding of equipotency ^9,10^.

The introduction of large language models (LLM) has revolutionized information technology. In the past few years, a number of LLMs have been introduced, showing proficiency in tasks ranging from summarizing information to text generation and data analysis. LLMs have shown potential for curating information more generally and participating in medical knowledge tasks more specifically ^11^. Models fine-tuned on medical texts can pass medical exams and aid clinical assessments ^12^. Although they have shown remarkable potential for medical knowledge and assessment, their real-world clinical utility remains unclear due to concerns over faulty clinical reasoning, diagnostic accuracy and deviation from standards-of-care, ^13^ particularly in the real-world setting ^14^ and in applications with the potential for bias. For medical decisions with potentially fatal consequences (such as opioid overdose), model performance, in terms of accuracy and reliability, is extremely important to evaluate and quantify. LLMs have been found to have challenges with arithmetic in conditions such as lengthier questions or more diverse mathematical operations ^15^.

In this study, we systematically-investigated how various commercially-available LLMs [AI21 (Jamba Large 1.7), Anthropic (Claude 3.5 Sonnet), Cohere (Command R+ 08-2024), DeepSeek (DeepSeek Chat), Google (Gemini 2.5 Flash, Gemini 2.5 Pro), Meta (Llama 3.3 70B Instruct), Mistral AI (Mistral Large 2407, Mixtral 8x22B Instruct), OpenAI (GPT-3.5 Turbo, GPT-4.1, GPT-4.1 Mini), and Qwen (Qwen 2.5 72B Instruct)], performed opioid conversions, with and without reference to a highly-cited paper which synthesized available clinical guidelines and studies to produce a comprehensive pharmacological reference ^16^. We sought to primarily determine whether LLMs, generally-trained, could safely and consistently provide opioid conversions within the context of clinical scenarios, and develop benchmarks for accuracy for commonly-performed drug conversions.

## Methods

### Ethical Considerations

This study did not involve patient data or human subjects. It was thus exempt from institutional review. Vignettes used in the study were fictional and not derived from specific patients. The Strengthening the Reporting of Observational Studies in Epidemiology (STROBE) reporting guidelines were followed for this investigation. Artificial intelligence was not used for experiment design, but Claude Sonnet 4.5 (Anthropic, San Francisco, CA) and GPT-4.1 (OpenAI, San Francisco, CA) were used at times to assist with code implementation.

### Study design and prompt framework

The entire experimental pipeline was implemented in Python 3.14.0 and is publicly available at https://github.com/tmarton1/opioid-conversion-experiment. In order to explore the variability of measurements between different chatbots, a number of commonly-used synthetic opioids were designated for conversion. Commercially-available LLMs were tested for accuracy and reliability of conversion (Table 1). In order to run simultaneous queries, an OpenRouter API (application programming interface; OpenRouter Inc. New York, NY) was called in order to control parameters and ensure that each query was independent and therefore not directly influencing the next query. This was used across all tested models using standard parameters (Table 1). In order to standardize queries across conditions and reduce variability, prompts were generated with a customized PromptService (PromptService) class. The framework for subsequent experiments is shown in Supplemental Figure 1.

**Table 1:** Details of large language models.

| Model Name | Developer | Year | Parameters (estimate) |
| --- | --- | --- | --- |
| GPT-4.1 | OpenAI | 2025 | 1.8 trillion |
| GPT-4.1 mini | OpenAI | 2025 | 8 billion |
| GPT-3.5 turbo | OpenAI | 2023 | 20 billion |
| Claude-3.5-sonnet | Anthropic | 2024 | undisclosed |
| Mistral Large 2 | Mistral AI | 2024 | 123 billion |
| Mixtral 8x22B-instruct | Mistral AI | 2024 | 39 billion active (of 141) |
| Gemini-2.5-pro | Google | 2025 | variable |
| Gemini-2.5-flash | Google | 2025 | variable |
| Command-r-plus-08-2024 | Cohere | 2024 | 104 billion |
| Jamba-large-1.7 | AI21 Labs | 2025 | 94 billion active |
| Qwen-2.5-7.2-instruct | Qwen (Alibaba) | 2024 | 72 billion |
| Llama-3.3-70b-instruct | Meta | 2024 | 70 billion |
| Deepseek-chat | Deepseek | 2025 | 671 billion |

### Data preparation

Raw data from a synthesis of published oral morphine equivalent (OME) conversion factors by Nielsen et al ^16^, a highly-cited synthesis of published OME doses from clinical guidelines and literature, were converted to Javascript Object Notation (JSON) format. Opioids of interest were selected by Y.C. and D.C. The first tier of opioids (in terms of importance) were oral morphine, oral hydromorphone, intravenous hydromorphone, intravenous morphine and oral oxycodone. Ten additional opioids were subsequently added for analysis, including oral codeine, oral hydrocodone, oral methadone, oral oxymorphone, oral tapentadol, oral tramadol, sublingual buprenorphine, transdermal fentanyl, intravenous fentanyl, and subcutaneous fentanyl. All opioids of interest were first converted to OME **(Table 2)** using the recommended median conversion from Nielsen et al ^16^. The API queried models independently. That dose of oral morphine was then converted into the corresponding dose of the “convert from” opioid, presented in units in Nielsen et al ^16^. For each request, we calculated the correct expected dose of the “convert to” opioid. We took the observed dose in the LLM’s response and calculated the conversion coefficient implicitly used by the LLM. We plotted the inferred coefficient as a function of the dose of the “convert from” opioid.

**Table 2:** Opioid conversions (from Nielsen et al)

| **Primary Opioid** | **Route** | **Recommended Conversion to OME (Median)** | **LQ-UQ** |
| --- | --- | --- | --- |
| **Morphine** | Oral (index) | 1 (mg/day) | 1-1 |
|  | IV | 3 (mg/day) | 3-3 |
| **Oxycodone** | Oral | 1.5 (mg/day) | 1.5-1.5 |
| **Hydromorphone** | Oral | 5 (mg/day) | 5-5.5 |
|  | IV | 17.5 (mg/day) | 16.9-18.1 |
| **Hydrocodone** | Oral | 1.2 (mg/day) | N/A |
| **Tramadol** | Oral | 0.2 (mg/day) | 0.2-0.2 |
| **Methadone** | Oral | 4.7 (mg/day) | N/A |
|  | Intravenous | 13.5 (mg/day) | N/A |
| **Buprenorphine** | Sublingual | 38.8 (mg/day) | 37.5-43.8 |
|  | Transdermal | 2.2 (mcg/hr) | 1.9-2.4 |
|  | Intramuscular | 75 (mg/day) | 75-90.6 |
|  | Intravenous | 75 (mg/day) | 75-90.6 |
| **Fentanyl** | Intravenous | 0.2 (mcg/day) | 0.2-0.3 |
|  | Transdermal | 2.7 (mcg/hr) | 2.4-3.2 |
|  | Subcutaneous | 0.2 (mcg/day) | 0.2-0.3 |
| **Nalbuphine** | Parenteral | 3 (mg/day) | 3-3 |
| **Oxymorphone** | Oral | 3 (mg/day) | N/A |
| **Codeine** | Oral | 0.1 (mg/day) | 0.1-0.2 |
| **Tapentadol** | Oral | 0.4 (mg/day) | 0.4-0.4 |

### Clinical scenario testing

In order to evaluate model performance in a real-world medical setting, clinical scenarios were tested in all models. These common clinical scenarios show situations where patients require changes in their opioid regimen, such as uncontrolled postoperative pain in patients on buprenorphine or patients with cancer and other multiple comorbidities. To account for the potential variability of effects such as wording, medication type, dosing schedule and patient-specific factors (age, demographics), we wrote 20 fictional scenarios which simulated real-life clinical encounters (see Supplemental Table 1). Scenarios were written by Y.C. and D.C. who have >20 combined years of clinical experience. Pharmacological appropriateness (e.g. availability of formulation in the United States) was further evaluated by a clinical pharmacist specialized in acute pain management (L.L.). In order to analyze the impact of wording, clinical scenarios were further revised into 4 variations (vignettes) using GPT 5.1 (OpenAI, San Francisco, CA). A fifth “empty” vignette was also created as a control (no scenario). Details of the prompt used for generating vignettes are available in Github (Microsoft, Redmond, WA) or upon request.

Performance was assessed using the same accuracy thresholds applied in the primary analysis: tight accuracy (0.85-1.15) and broad accuracy (0.6-1.7). We chose these thresholds based upon published recommendations for opioid weaning decrements to avoid withdrawal symptoms (ranging from 10-20% per time period) ^17,18^, and on opioid uptitration (specifically in patients with cancerr, a scenario where dose increases may be required more aggressively) ^19^. Results were stored in CSV format with nested arrays containing all observed doses for each model under each prompting condition, enabling analysis of both central tendency and response variability across repeated trials. For scenarios requiring reductions for incomplete cross-tolerance, multipliers (e.g. multiply by 0.75) were built into code to calculate the correct equipotent conversion. P values reflect either 1) tight performance if significant or 2) broad performance if tight performance is not significant, between adjacent models for comparison.

### Opioid pair random dose conversions

In order to further elucidate sources of response variability and inaccuracy, random doses of opioids were also tested. For each API request, expected conversion values were calculated with published equianalgesic conversion factors from a JSON describing the reference dataset and stored alongside the request time, model used, source and target opioid details, the full prompt sent to the model, boolean prompt descriptors, the raw model response, and the regex-extracted model-output dose in comma-separated values (CSV) results table format ^16^. Opioid pairs were ranked by clinicians (Y.C. and D.C., who have board certification in pain medicine and fellowship training in regional anesthesiology and acute pain medicine, respectively) based on clinical relevance. Lower priority opioids were each converted into daily oral morphine equivalents; highest priority pairs were tested in full permutations in both conversion directions. For each conversion pair, 25 independent trials were carried out, using a log-uniform random value for the input dose, which ranged from the equivalent of 10-120 milligrams per day of oral morphine (for statistical robustness and to mirror real-life clinical ranges, such as Washington State Health Care Authority, which limits to 120 MME daily without consultation with pain specialist and other state guidelines) ^20,21^.

### Rescue architecture

In order to investigate the effects of prompt augmentation, a prompt was constructed to include conversion values from Nielsen et al. in JSON format and processed with Python 3.14.0. JSON data were further converted into CSV format for subsequent data analysis, permitting reference-based prompting (or prompt augmentation) to improve model accuracy. Sequential prompts were used to improve the accuracy of tested foundational models.

### Statistical analysis and visualizations

All descriptive statistics were computed using statsmodels v0.14.6, SCIPY v1.16.3, pandas 2.3.3, and NumPy 2.3.4 packages in Python. Visualizations made with Bokeh 3.8.1, a Python-based visualization library. For model ranking experiments, the Cochran–Mantel–Haenszel test was used to compare models accuracy across the different strata (tight and broad accuracy), and Bonferroni corrected. Models without significant differences were given the same rank. Effects of vignette text, explaining units, and including the Nielson reference were measured using Fisher’s Exact Test and Bonferroni corrected. Pearson’s χ2 tests of independence were used to test the relationship between model accuracy and response precision (coefficient of variation) when grouped into 3 x 3 contingency grids. Bonferroni-corrected Mann–Whitney U tests were used to test pairs of vignettes wordings where response distributions differed significantly in model-scenario cells. Wording effects were also investigated using intraclass correlation analysis. An ANOVA decomposed the total variance into between-vignette and within-vignette variance, from which the intraclass correlation coefficient (ICC) metric and F-test were calculated. Then separated Spearman’s rank correlation *ρ* was used to test whether scenarios with higher total response variance also had higher intraclass correlation coefficients. The overall distribution of Spearman’s *ρ* values were assessed with a binomial sign test. Kendall’s coefficient of concordance (W) was used to test model agreement regarding vignette wordings and higher or lower responses within each scenario. To test prompt augmentation effects, Wilcoxon signed-rank test was used to see if model accuracy changed on conversion items (and also for differences in summative analysis for naive models). Code available on Github (Microsoft, Redmond, WA) or upon request.

### Safety and adverse events

Real patients were not included in this study. There were no events of any type and no concerns with data security.

## Results

### LLM safety in clinical performance

In naive conditions, LLMs achieved low accuracy in performing conversions required in clinical vignettes (Figure 1A, ranked performance). Tight-range accuracy (0.85-1.15 fold) was achieved with 16.3-41.1% of model conversions, while broad-range accuracy (0.6-1.7 fold) was achieved between 49.4-82.9% of the time by models. Interestingly, models would often provide the wrong conversion consistently (Figure 1B), with coefficient of variation (CV) < 0.4 for the majority of clinical scenarios, showing consistency even in inaccurate answers (median CV = 0 for all models except DeepSeek-Chat). Figure 1C ranks models by the distribution of vignette conversions with tight accuracy, incorrect but precise conversions, and imprecise and inaccurate conversions. Accuracy distribution is also shown by CV “bin,” where notably most modes did not contain significant numbers of observations with CV>0.2, with the exceptions of DeepSeek-Chat and Gemini-2.5-Pro (Figure 1D).

**Figure 1:**
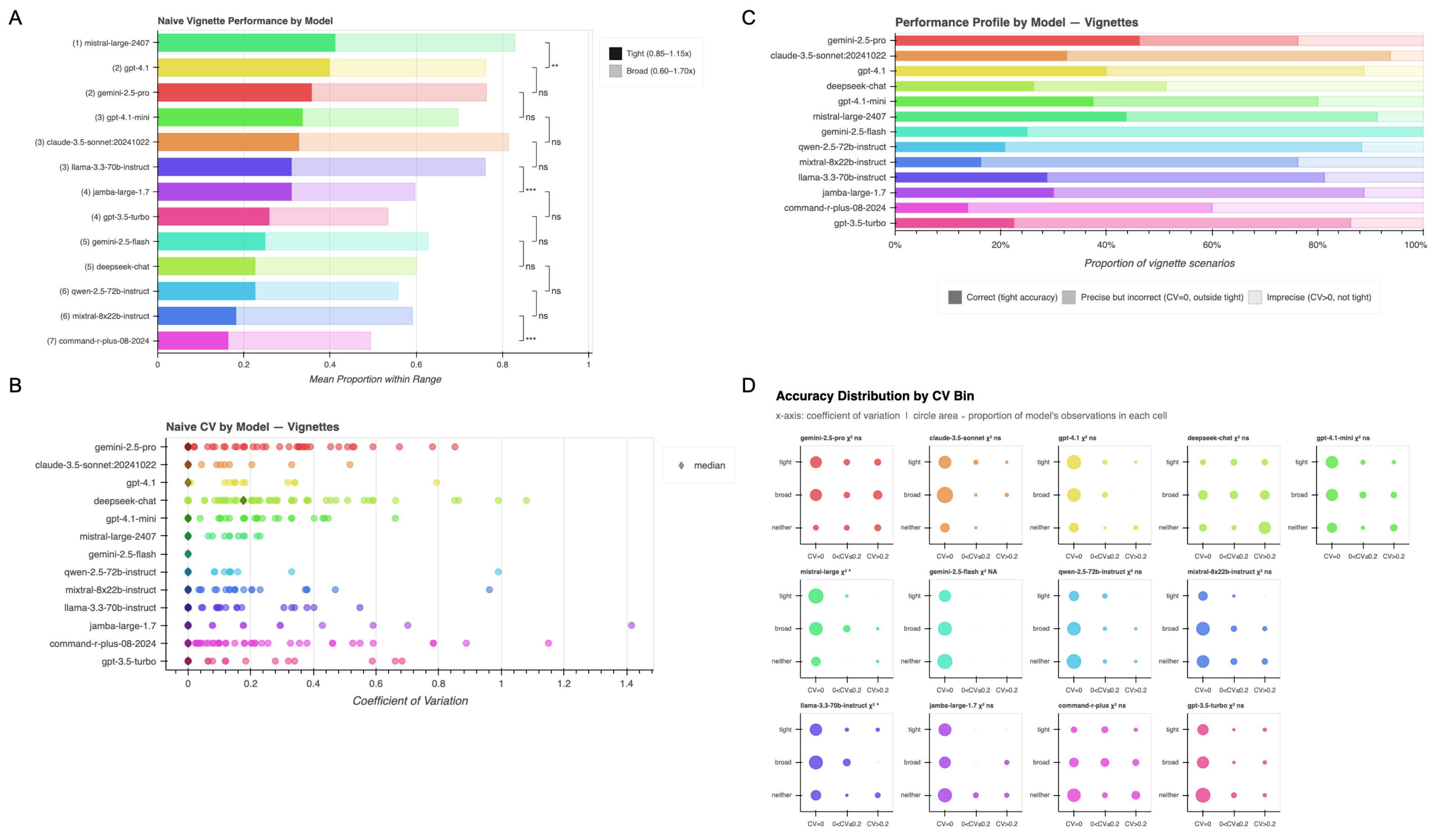
**A)** Horizontal bar chart showing the proportion of model responses falling within the tight (solid bar, 0.85–1.15x) and broad (translucent bar, 0.6–1.7x) accuracy ranges for vignette-based opioid conversion prompts, without any reference information (naive). Models are ordered by global rank. The significance brackets compare adjacent-ranked models with Cochran–Mantel–Haenszel tests (p values indicated in figure). **B)** Horizontal strip charts showing coefficient of variation (CV) for each vignette (with 7 repetitions). Diamond represents the median CV. **C)** A bar graph showing proportion of conversions in vignettes with tight accuracy, which were precise but inaccurate, and imprecise and inaccurate. **D)** Accuracy “bins” plot accuracy and CV for each model; circle size is proportional to number of observations in each category. ***p<0.001. **p<0.01. *p<0.05

### Response variance across models

In order to investigate the sources of variance between vignettes (the wording of the clinical situation), scenarios were re-written into four variations (multiple vignettes). Across a total of 28 responses (7 trials per vignette variation), higher accuracy necessarily limited variability. Between-vignette variability is much more notable (Figure 2A and B), and is attributed to the effect of wording, rather than stochastic noise (which would be equally distributed between secondary vignettes from the same clinical scenario). Overall, 76% of variance occurs between vignettes across the same scenario. Scenarios with high total variance showed higher intraclass correlation (ICC), suggesting that sensitivity to vignette wording corresponds to high response variability, Spearman’s *ρ* = 0.375, p < 0.001 (Figure 2C). This was specifically true for mistral-large-2407 (*ρ* = 0.57, p = 0.009), gpt-4.1 (*ρ* = 0.54, p < 0.014), claude-3.5-sonnet:20241022 (*ρ* = 0.64, p = 0.002), llama-3.3-70b-instruct (*ρ* = 0.70, p < 0.001), gemini-2.5-flash (*ρ* = 0.83, p < 0.001), qwen-2.5-72b-instruct (*ρ* = 0.62, p = 0.004), mixtral-8x22b-instruct (*ρ* = 0.76, p < 0.001); notably gemini-2.5-pro did not exhibit this variability (*ρ* = 0.05, p < 0.850) (Figure 2C).

**Figure 2:**
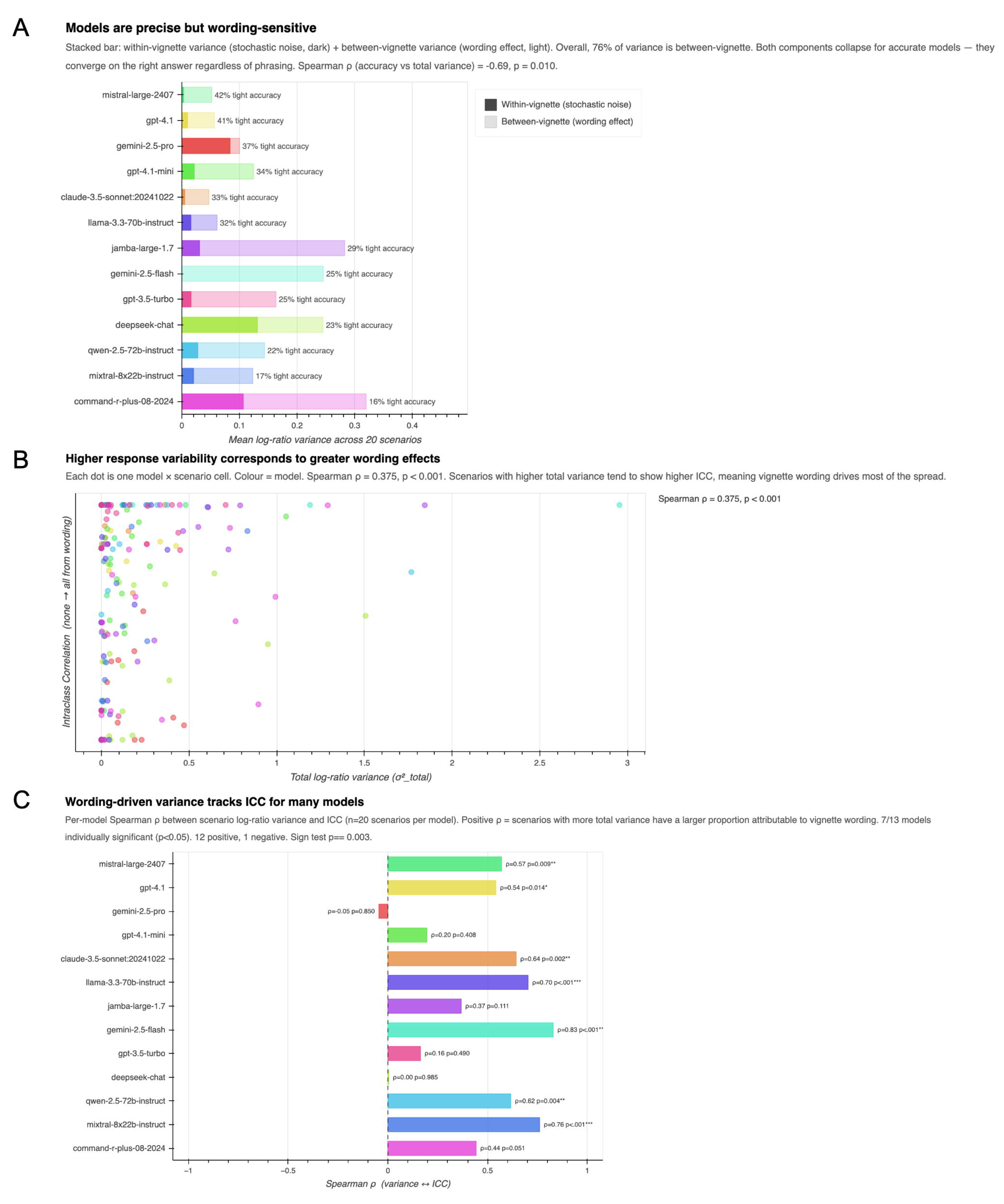
**A)** Horizontal bar graph showing mean scenario log-ratio variance. Solid bar shows the proportion of variance attributable to within-vignette variability measured from 7 observations per model; translucent bar shows the proportion of variance attributable to between-vignette variability measured across 4 vignettes per scenario. Models are ranked by accuracy. B) Total scenario variance is plotted against intraclass correlation. Each dot represents one model by scenario. Model-scenarios with higher variance tend to attribute variability to vignette wording. C) Horizontal bar graph showing Spearman’s rank correlation (ρ) by model, between scenario log-ratio variance and ICC. Models with positive ρ exhibit higher proportion of variance attributable to vignette wording.

When performing pairwise comparisons between vignettes (6 possible combinations), a large number of scenarios exhibited statistically-significant differences between vignettes (Figure 3A). In order to ascertain whether models “agree” or “disagree” on wording effects, Kendall’s coefficient of concordance (W) was used (Figure 3B), showing low concordance and thus the lack of systematic cross-model source of wording effects.

**Figure 3:**
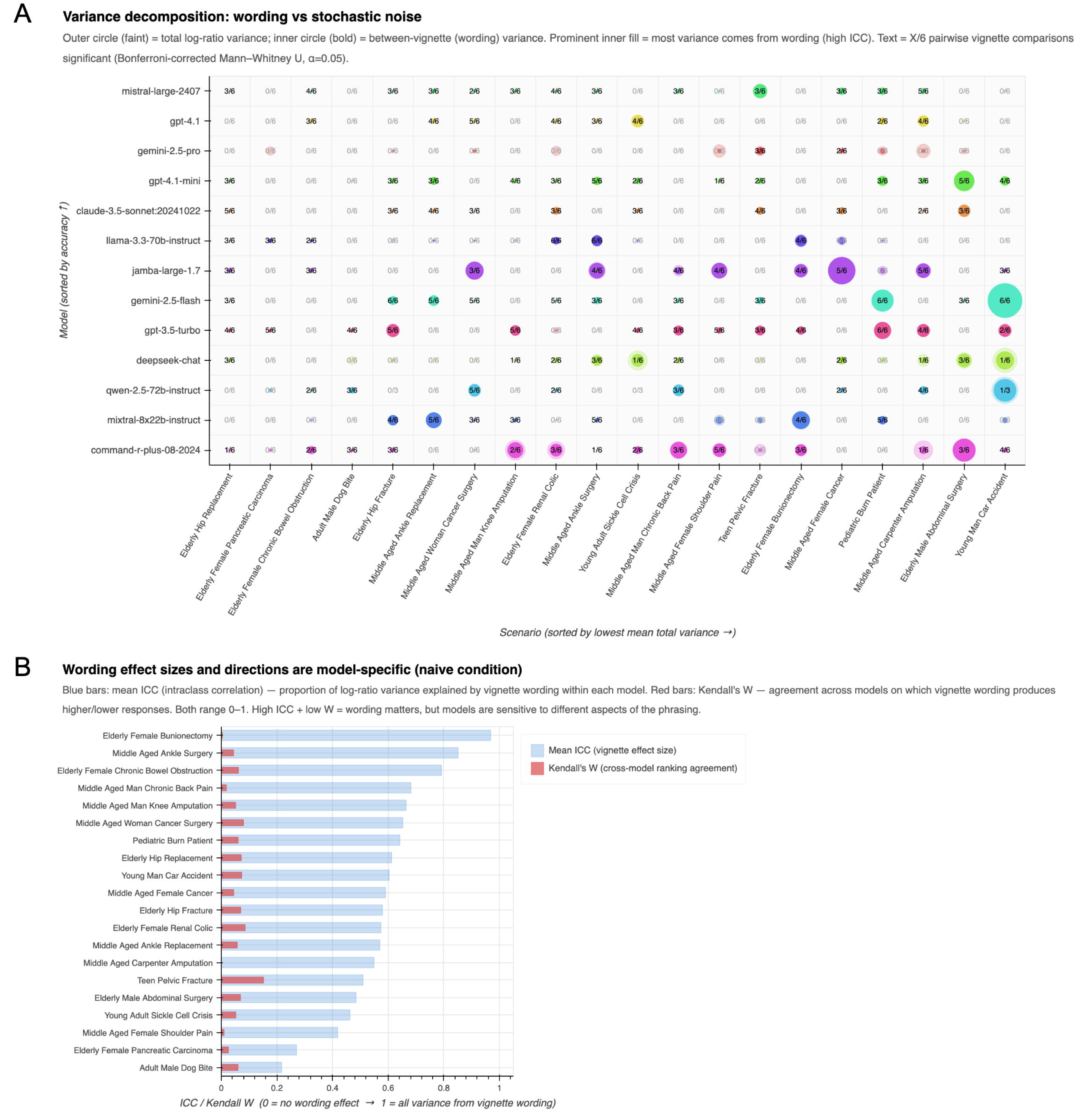
**A)** Concentric circle grid plotting model by clinical scenario, sorted by naive model accuracy in vignette conversion. The translucent outer circle area corresponds to total model-scenario log-ratio variance while the opaque inner circle area indicates between-vignette variance (indicating effects of wording). “X/6” indicates the proportion of vignette:vignette comparisons which are statistically-significantly different from one another after Bonferroni correction, for each model-scenario. **B)** Paired bar chart showing ICC/Kendall W for each scenario. Blue bar indicates the mean ICC, or proportion of response variance attributable to wording effects within each model. Red bar indicates the Kendall’s W, or the cross-model agreement on which working yields higher or lower responses.

### Random dose conversions

Without the context of a clinical scenario, model ranking mirrors the prior vignette results (random-dose opioid pairs [n=49]) (Spearman’s *ρ* = 0.67). Figure 4A shows ranked performance between models (p values reflect either 1) tight performance if significant or 2) broad performance if tight performance is not significant, between adjacent models). CV was more randomly distributed in this experiment (Figure 4B), yet all median CV values were <0.5.

**Figure 4:**
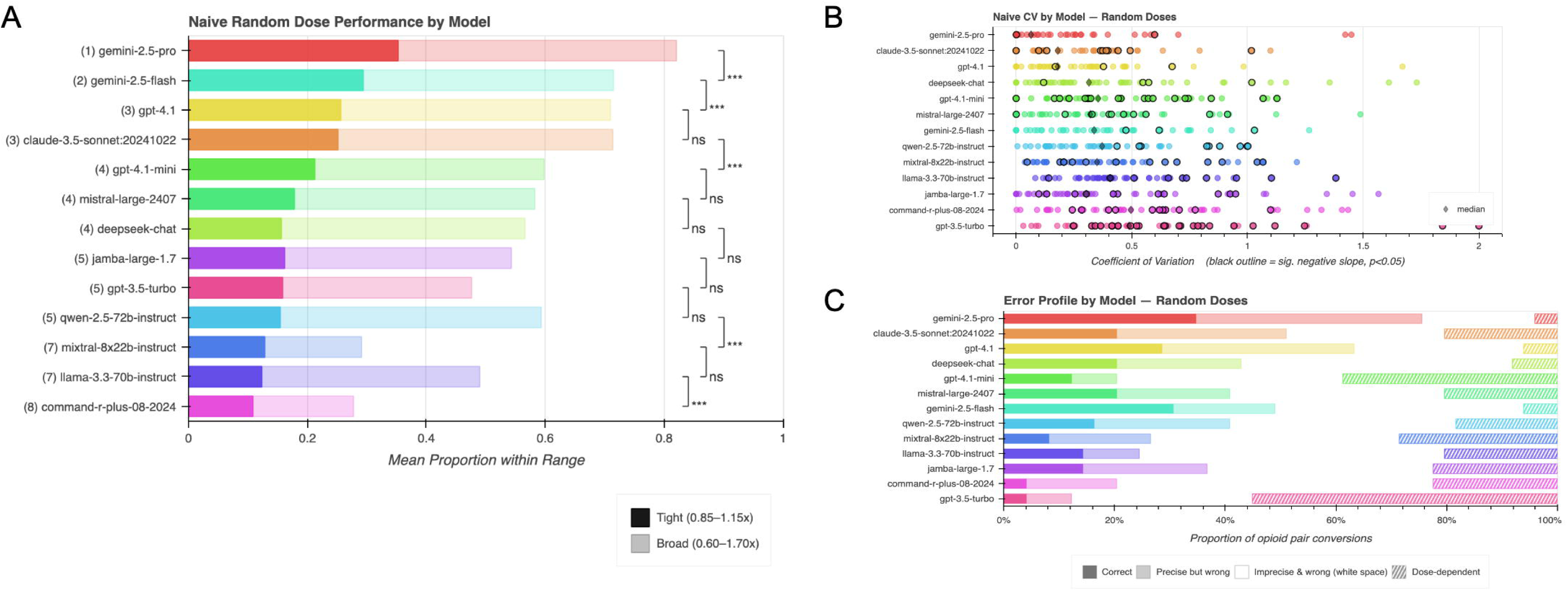
**A)** Horizontal bar chart showing the proportion of model responses falling within the tight (solid bar, 0.85–1.15x) and broad (translucent bar, 0.6–1.7x) accuracy ranges for controlled random dose conversions, without any reference information (naive). Models are ordered by global rank. The significance brackets compare adjacent-ranked models with Cochran–Mantel–Haenszel tests (p values indicated in figure). **B)** Horizontal strip charts showing coefficient of variation (CV) for each opioid pair (with 25 random dose repetitions per pair). Diamond represents the median CV. **C)** Bar graph shows the proportions of types of error by model tested. On the right, the proportion of opioid pairs where the inferred coefficient of conversion and the expected dose had a statistically significant (p<0.05) negative slope (dose-dependent), indicating a failure to properly deploy a ratio calculation. All other opioid pairs exhibited a near-zero slope between the coefficient of conversion and the expected dose, and were either correct, incorrect but precise (CV < 0.2), or incorrect and noisy.

Figure 4C shows that the majority of incorrect doses did not show a dose-dependent conversion coefficient, suggesting the application of a consistent (albeit wrong or noisy) conversion coefficient. However, a proportion of opioid pairs did display a slope statistically-significantly different from zero, indicating an inability to perform a consistent conversion ratio (notably for gpt-3.5-turbo) (see cross-hatched bars in Figure 4C).

### Summative analysis for naive performance

Performance in naive models was similar across multiple experimental paradigms (random-dose “pair” conversions, empty vignettes and text-based real world vignettes restricted to 11 opioid pairs in all conditions, Figure 5). Wilcoxon signed-rank tests on broad-range accuracy showed statistically-insignificant differences, with the exception of llama-33-70b and command-plus-08-2024 (p<0.05).

**Figure 5:**
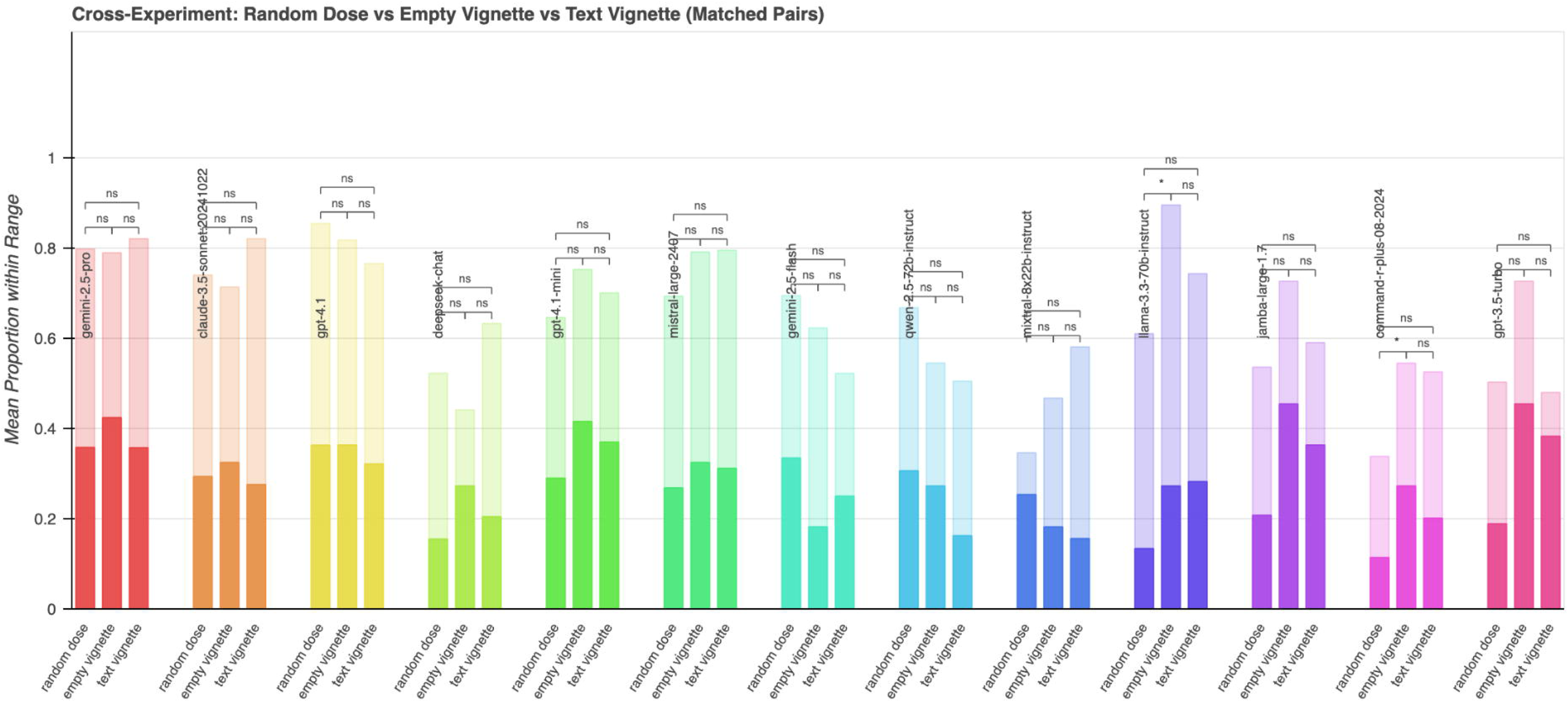
Grouped bar graph compares each model across 3 experimental paradigms: random-dose opioid-pair conversions, “empty vignettes” (without patient text), and text-based vignettes, standardized by the 11 opioid pairs common to these three paradigms. Significance brackets show statistically-significant differences in broad-range accuracy by paired Wilcoxon signed-rank test.

### Prompt augmentation improves performance

Sequential prompt augmentation was used to improve model performance. Incorporation of a reference table in prompting (from Nielsen et al ^16^) improved performance significantly in clinical vignettes and randomly-generated dose pair-wise conversions. Wilcoxon signed-rank test showed statistical-significance (p<0.001-0.05) in 11 of 13 models for clinical vignettes (Figure 6A), and all models for random doses (p<0.001). (Figure 6B). A closer examination of reference augmentation showed that the “Nielsen effect” was significantly more pronounced in conversions to oral morphine (OME) (Figure 7), compared with other synthetic opioids. For vignette conversions, 7 models showed highly statistically-significant differences with Nielsen et al ^16^, converting to OMEs (p<0.001), while none displayed this for non-OME conversions. This was more notable for the random dose experiment.

**Figure 6:**
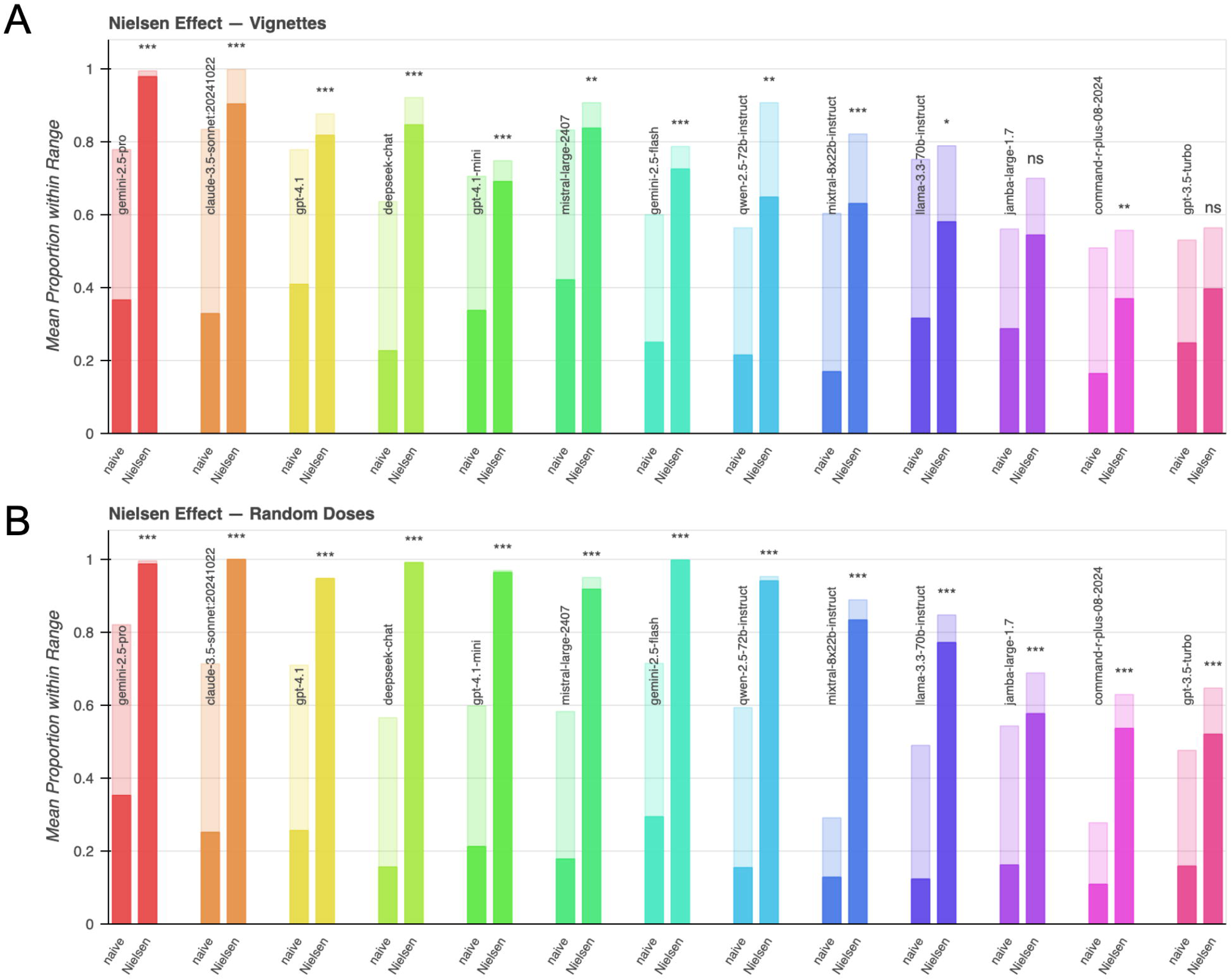
Horizontal bar graph shows how prompt augmentation with the Nielsen grounding reference impacts broad and tight accuracies in **A)** vignettes and **B)** random dose experiments. Wilcoxon signed-rank test indicates statistically-significant differences between models.

**Figure 7:**
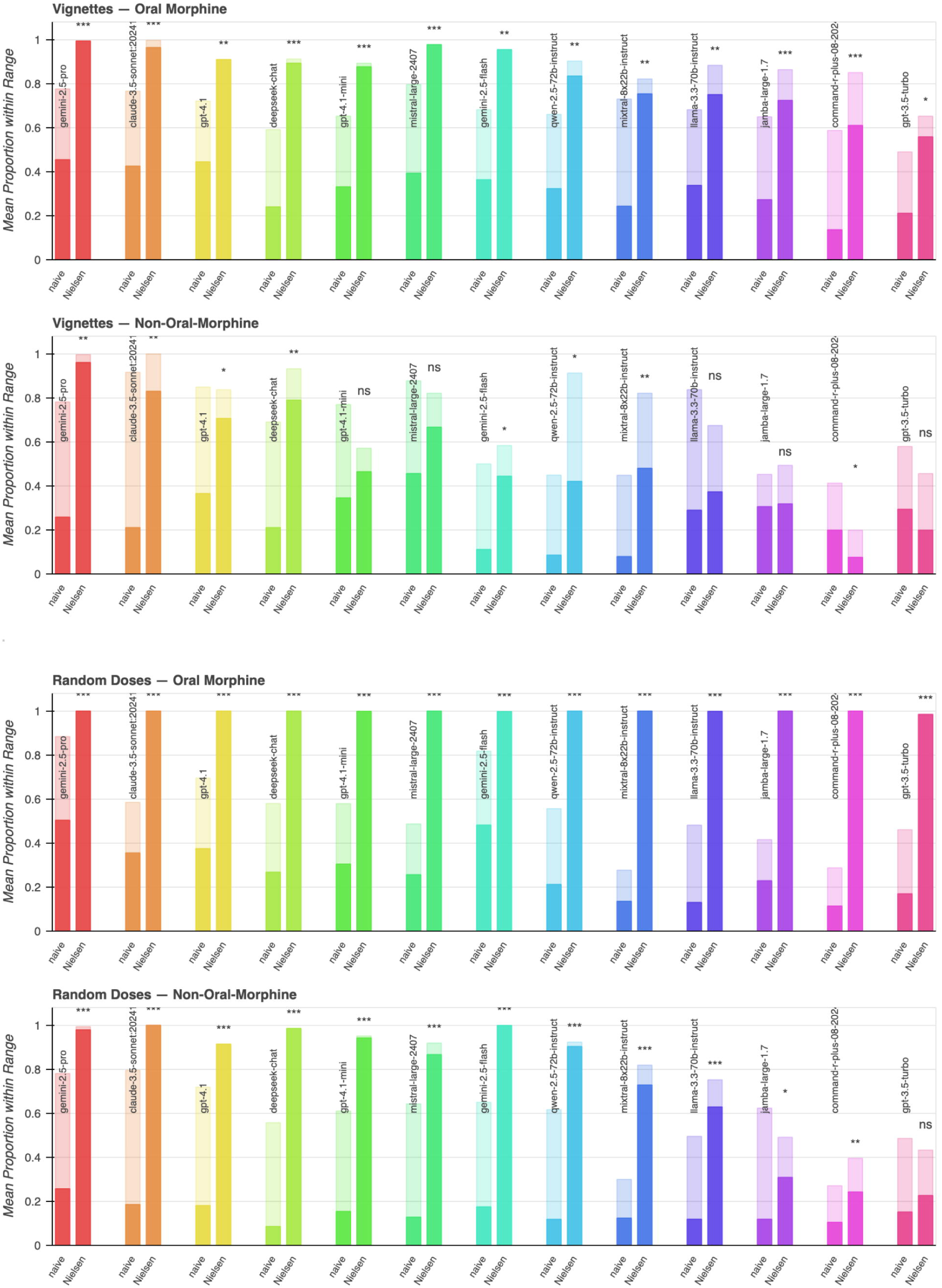
Horizontal bar graphs showing “Nielsen” effect by conversion type (opioid to OME versus opioid to opioid), and experiment. Significance brackets show statistically-significant differences in broad-range accuracy by paired Wilcoxon signed-rank test. ***p<0.001. **p<0.01. *p<0.05

Notably, non-daily dosing (e.g. dose every 4 hours or three times a day) was found to confuse models when conversions to oral morphine (OMEs) were requested for a different unit of time (every 4 hours) (Figure 8A). Unit explanation incorporated in the prompt further produced significantly-improved conversions across models (Figure 8B), notably for gpt-4.1, deepseek-chat, gpt4.1-mini, mistral-large-2407, qwen-2.5-72b-instruct, mixtral-8x22b instruct), and proved more effective than reference-grounding alone in improving performance for several models (claude-3.5-sonnet:20241022, gpt-4.1, deepseek-chat, gpt-4.1-mini, mistral-large-2407, qwen-2.5-72b-instruct).

**Figure 8:**
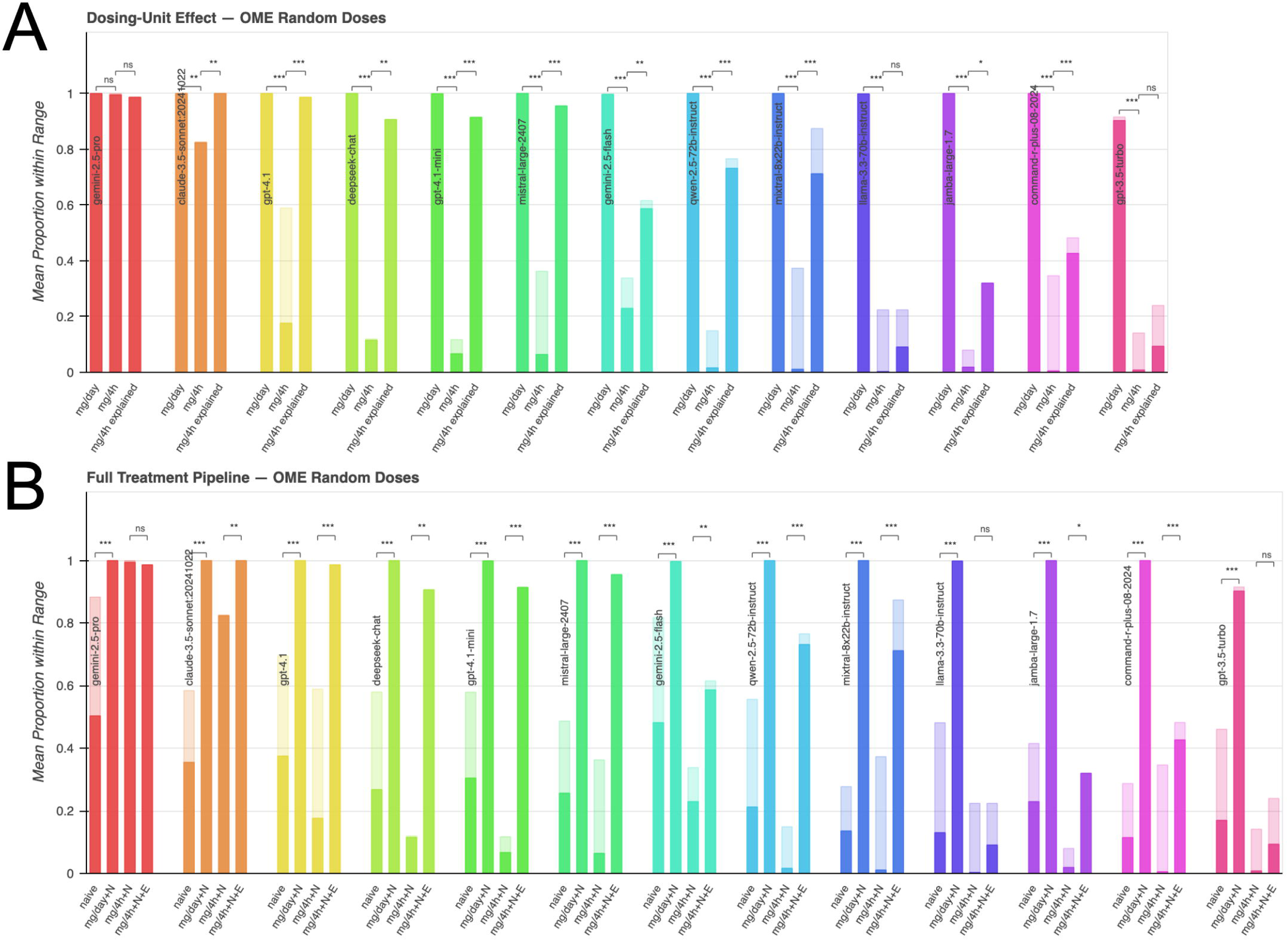
**A)** Barcharts show the effect of unit explanations in the random-dose experiments in which all opioids were converted to oral morphine in either mg/day or mg/4h units. The Wilcoxon signed-rank statistical significance of the accuracy distribution difference across units is annotated. For mg/4h prompts, the effect of adding a paragraph calling attention to and explaining the units is compared to withholding that paragraph. The statistical significance of adding the explanatory paragraph was measured by Wilcoxon signed-rank test. B) The Wilcoxon signed-rank statistical significance of adding the Nielsen reference to mg/day oral morphine conversions is annotated alongside the significance of adding the paragraph explaining units to the mg/4h prompts.

The combined effects of unit explanation and reference grounding in prompt augmentation significantly improved tight accuracy in models performing both random-dose exercise conversions (for OME conversions) and vignette conversions (Figure 9). After including these modifications, 6 of the 13 models achieved >90% tight-range accuracy on non-daily oral morphine conversions and 2 of the 13 models achieved >90% accuracy on the vignettes, which include 2 non-daily dosing scenarios (Figure 9). Overall, prompt augmentation improved the performance of larger models (gemini-2.5-pro, claude-3.5-sonnet:20241022, gpt-4.1) significantly, allowing >80% tight-range accuracy on vignettes.

**Figure 9:**
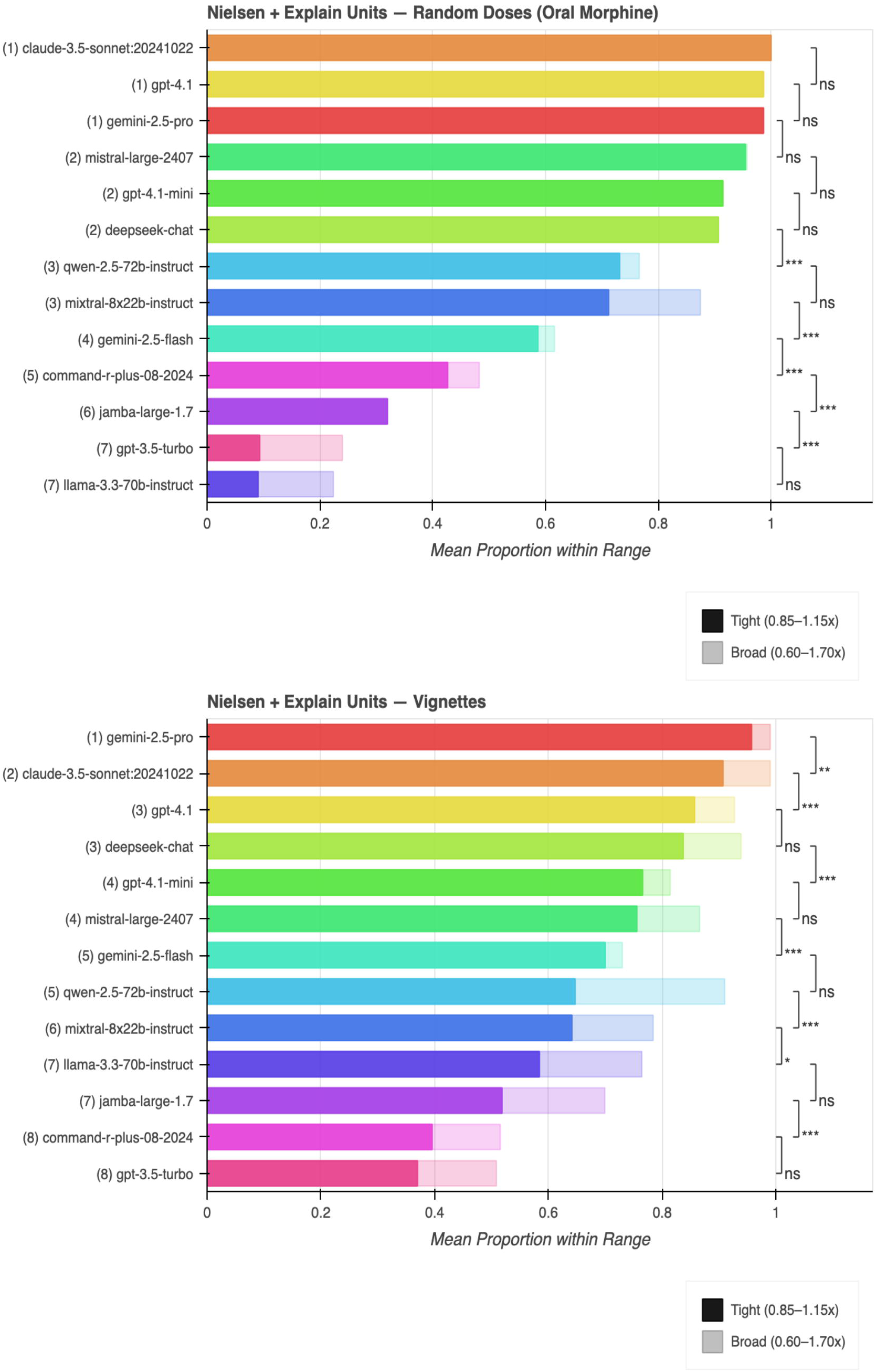
Top and bottom panels show best possible performance ranked by model, combining prompt augmentation methods (reference table, unit explanation) to improve performance in random doses pairs (to OME) and vignettes. The significance brackets compare adjacent-ranked models with Cochran–Mantel–Haenszel tests (p values indicated in figure).

## Discussion

In this systematic, API-based study, we investigated how a series of commercially-available LLMs performed in providing conversions of synthetic opioids (to morphine equivalents) and to equipotent doses of different opioids. In naive conditions, the vast majority of LLMs were inaccurate and at times, inconsistent across queries, failing to meet broad accuracy criteria in conversions. Alternatively, some models tended to consistently perform the same inaccurate conversions with low variability. These findings seem to be consistent with prior observations that LLMs perform relatively poorly at mathematics, despite recent advances in improving performance with strategies such as tool-calling, even for clinical tasks ^22^. They underscore the importance of caution when viewing the growing use of LLMs in medical education and clinical workflows.

Beyond demonstrating inconsistent performance, this study revealed the sensitivity of models to prompt wording. Re-wording of equivalent vignettes (for a particular scenario) accounted for 76% of variability within a particular scenario, exceeding what would be expected from stochastic sampling. This observation directly impacts the way in which clinicians interact with LLMs; narrative structure may significantly alter a model’s response and suggested course of treatment.

Opioid overdoses can occur in a variety of settings, including tapering or discontinuation ^23^, and even rotations ^24^. Thus, it is vital for reference sources to provide accurate equipotent conversions for patient safety. Language models predict words in sequence, excelling in tasks such as summarization or answering questions. Modern commercial LLMs make predictions of text based upon probabilistic relationships between words, informed by training on large corpora of data ^25^. Due to the randomness of statistical sampling, they may perform numerical calculations with significant variation. Our results suggest that without contextual prompting, LLMs may struggle to provide safe and consistent recommendations in clinical scenarios with narrow margins for error (medications with a narrow therapeutic index) and necessity for consistency. Thus, they pose risks from the perspective of safety in opioid stewardship and possibly other classes of medications with similar pharmacological action but different affinities (e.g. selective serotonin reuptake inhibitors). While LLMs are not fully-independent in healthcare, they are extensively used by trainees for medical school coursework ^26^ or clinical work ^27^, and thus potentially play a formative role in medical education.

Interpretability, or the ability to determine how LLMs reach conclusions or make decisions, is a crucial consideration in healthcare. Investigators have used strategies such as prompt engineering or chain-of-thought to better simulate clinical reasoning ^28^, permitting models to provide rationale for different decisions. Such approaches may increase trust and reduce the risk of errors due to hallucinations or other reasoning errors. In this study, we discovered that simple computational tasks involving drug conversions could be improved through prompt-engineering and context augmentation. Accuracy is vital for safe clinical care and it is vital for LLMs to avoid hallucinations, failures in logic, and behaviors such as sycophancy ^29^. Opioids are a commonly-prescribed medication with a relatively-high side effect profile. Thus, they are an important test case for LLM safety.

Strengths of this study include the formalized use of an API for scale (13 models tested) and independence of queries, breadth of opioids tested, repetition of queries to test reliability and novel approach to benchmarking performance. These measures allowed for a more comprehensive approach to assessment of model performance, particularly important to the rigor and evidence required for clinical therapy. An API permitted us to write independent queries, preventing chatbots from utilizing information from prior sessions in subsequent answers ^30^ and reducing inconsistency. Our testing platform also allowed an upscaled investigation of >15 models and multiple combinations of opioids/routes simultaneously, therefore yielding greater insight into patterns in errors and overall model safety. Overall, the rigor of our prompting permitted 1) consistency between queries, 2) statistical robustness and dose randomization and 3) analysis of the underlying operation performed by the language model in answering the prompt. Furthermore, the effect of variance specifically due to vignette wording reveals novel insights about the source of inaccuracy in naive LLM function.

Limitations to this study include the lack of real clinical data availability for model training. Currently, there is no definitive mechanism for ensuring privacy of protected health information (PHI) in commercial LLMs, which absorb information and thus cannot be used without caution. The authors’ institutions do not currently grant IRB approval for investigation of LLMs with PHI. While the authors attempted to simulate real-life clinical scenarios where opioids would require rotation, they may not capture the complexities (or chaos) of an actual patient encounter and electronic health record documentation. We also acknowledge the variability in opioid equivalence conversions in the literature ^5^, a factor which may have contributed to the inconsistency in conversion factors and is an important caveat to our use of a single reference for grounding (although this reference does synthesize multiple sources of data and is widely cited). Other weaknesses of this study include questions of generalizability; it is not certain whether our knowledge-grounding method may rescue performance for other types of agents.

In conclusion, we found that LLMs are insufficiently accurate for arithmetic-based opioid conversions in a naive setting, both in regards to simulated clinical scenarios and randomly-generated simple doses. Upon closer examination, many models consistently outputted the same inaccurate dosing across trials and in random dosing experiments, showing the ability to consistently apply the same, albeit incorrect ratio of conversion. Interestingly, clinical vignette wording often influenced the response and explained far more of the within-scenario variability than random noise. For most models, including the Nielsen conversion ratios to mg/day oral morphine in the prompt greatly improved accuracy, such that it was near-perfect across all investigated conversions to oral morphine. The ability to convert to non-oral morphine opioids was more variable across models, with larger models such as Gemini, Claude, and ChatGPT families performing best.

## Data Availability

All data produced in the present study are available upon reasonable request to the authors

---

**Supplementary Table 1:**
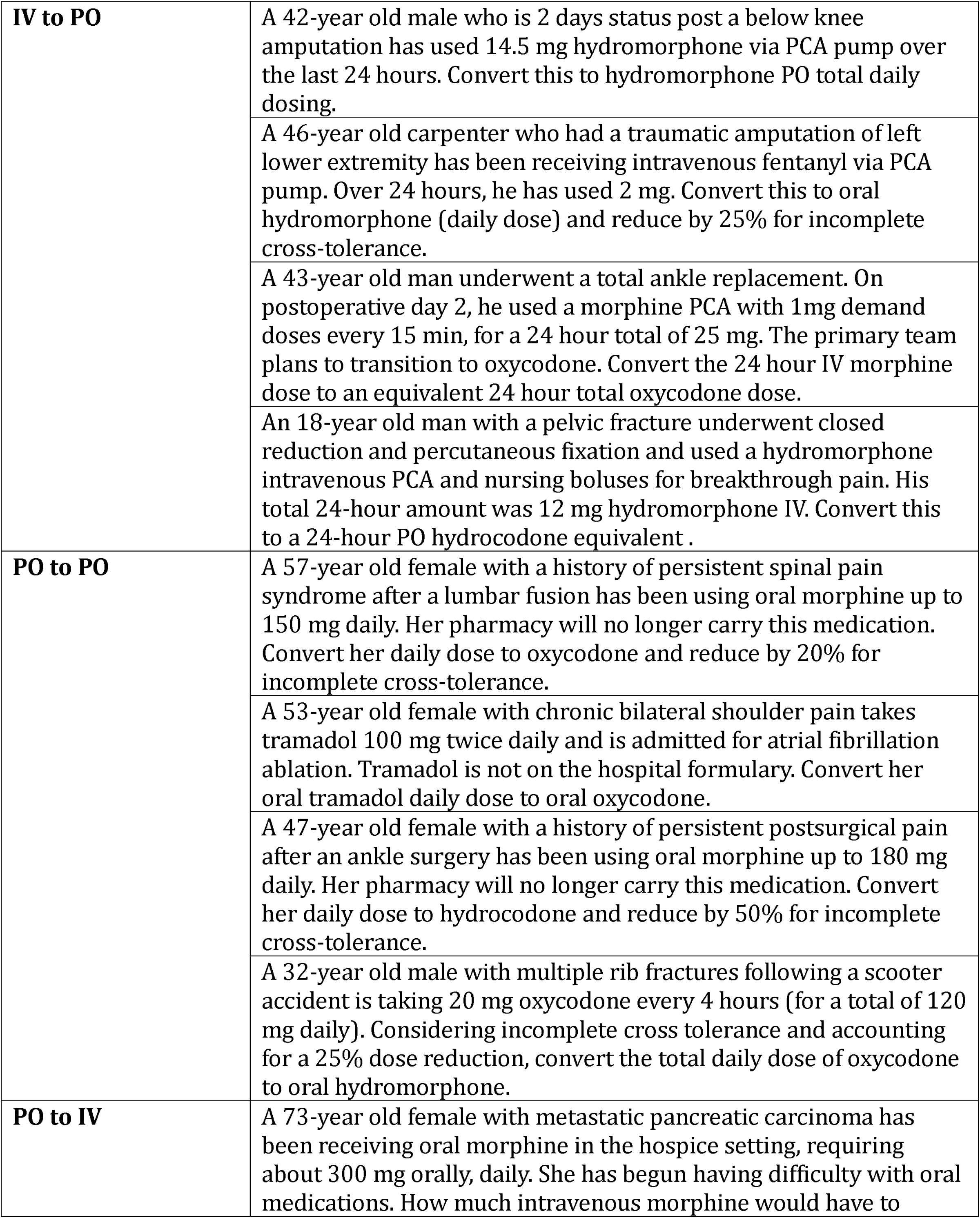
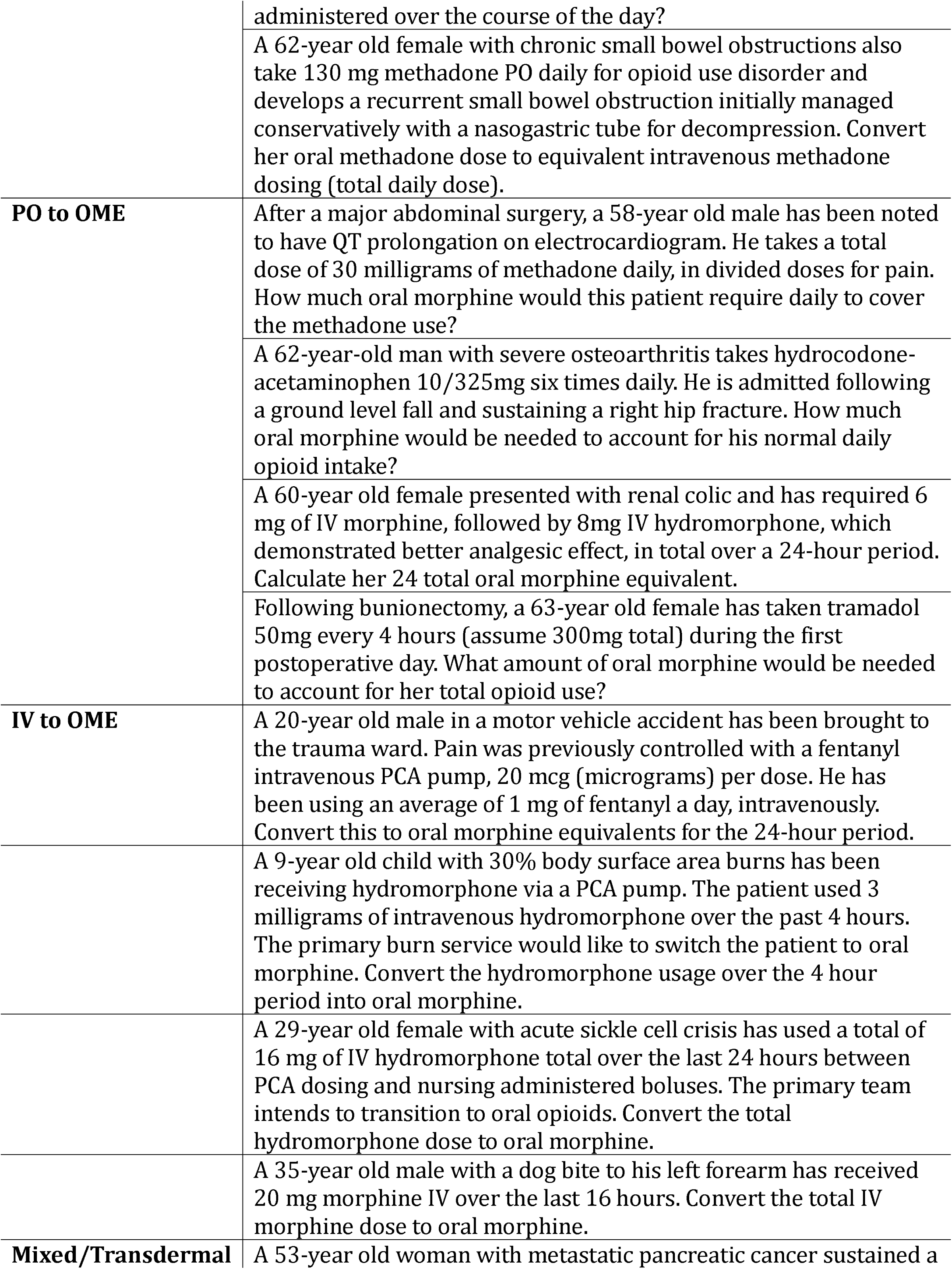
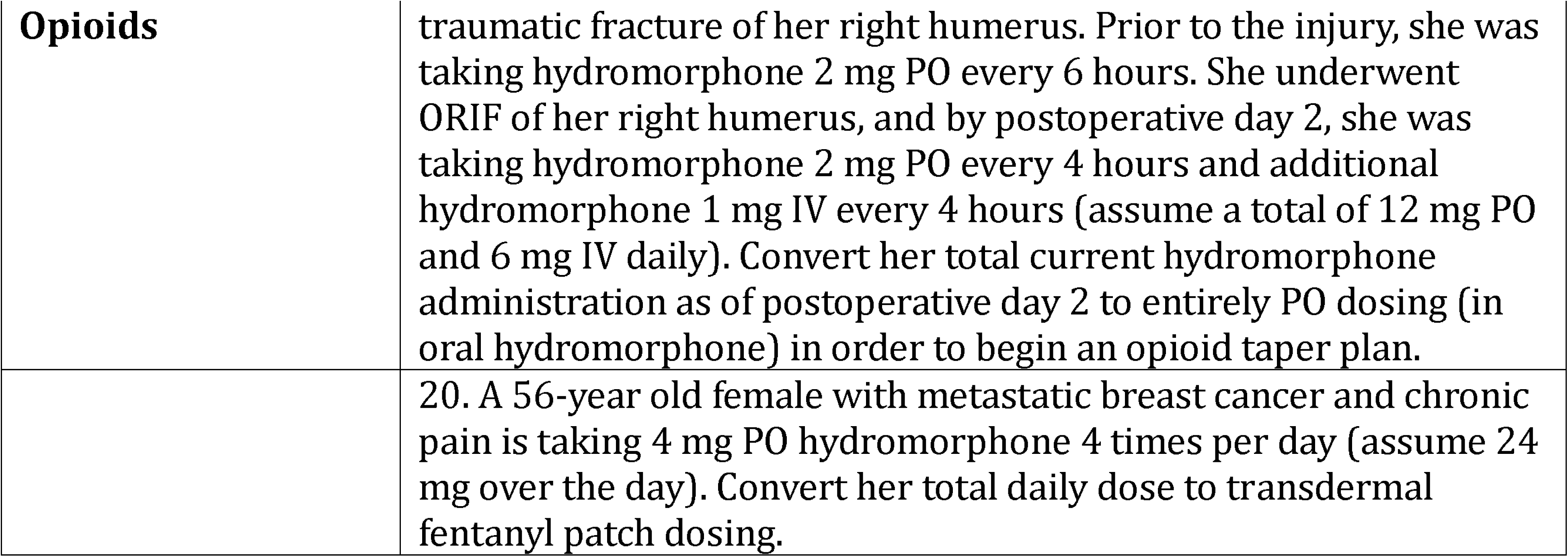
Vignettes.

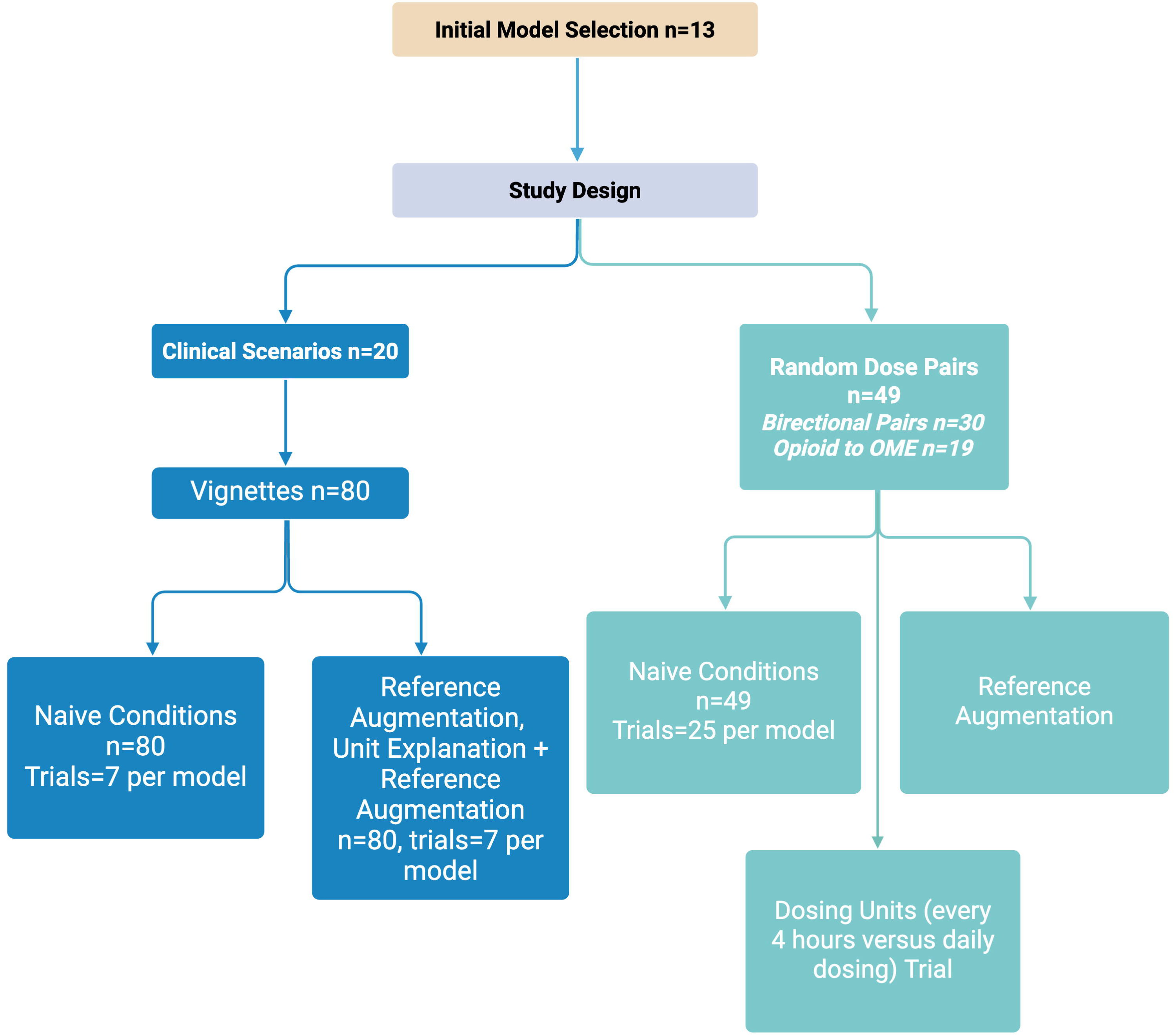

## References

1. Schoenfeld, A. J. et al. Reductions in sustained prescription opioid use within the US between 2017 and 2021. Sci. Rep. 14, 1432 (2024).

2. Khan, A., Irshad, M., Javed, Z., Naz, Z. & Fatima, M. Beyond opioids: FDA-approved suzetrigine offers hope for acute pain management. Ann. Med. Surg. (Lond.) 87, 4023–4025 (2025).

3. Treillet, E., Laurent, S. & Hadjiat, Y. Practical management of opioid rotation and equianalgesia. J. Pain Res. 11, 2587–2601 (2018).

4. Reddy, A. et al. Opioid rotation and conversion ratios used by palliative care professionals: An international survey. J. Palliat. Med. 25, 1557–1562 (2022).

5. Rennick, A. et al. Variability in opioid equivalence calculations. Pain Med. 17, 892–898 (2016).

6. Chaitowitz, M., Tester, W. & Eiger, G. Use of a comprehensive survey as a first step in addressing clinical competence of physicians-in-training in the management of pain. J. Opioid Manag. 1, 98–108 (2005).

7. Plagge, H. et al. Dose calculation in opioid rotation: electronic calculator vs. manual calculation. Int. J. Clin. Pharm. 33, 25–32 (2011).

8. Rurup, M. L. et al. The use of opioids at the end of life: the knowledge level of Dutch physicians as a potential barrier to effective pain management. BMC Palliat. Care 9, 23 (2010).

9. Hanks, S. The law of unintended consequences: when pain management leads to medication errors. P T 33, 420–425 (2008).

10. Vl Corcoran, R. Modes of administration of morphine in cancer pain. Some doctors lack basic knowledge about prescribing morphine. Br Med J 313, (1996).

11. Rao, A. S. et al. Large language model performance and clinical reasoning tasks. *JAMA Netw*. Open 9, e264003 (2026).

12. Singhal, K. et al. Toward expert-level medical question answering with large language models. Nat. Med. 31, 943–950 (2025).

13. Hager, P. et al. Evaluation and mitigation of the limitations of large language models in clinical decision-making. Nat. Med. 30, 2613–2622 (2024).

14. Bean, A. M. et al. Reliability of LLMs as medical assistants for the general public: a randomized preregistered study. Nat. Med. 32, 609–615 (2026).

15. Srivatsa, K. A. & Kochmar, E. What makes math word problems challenging for LLMs? in Findings of the Association for Computational Linguistics: NAACL 2024 (Association for Computational Linguistics, Stroudsburg, PA, USA, 2024). doi:10.18653/v1/2024.findings-naacl.72.

16. Nielsen, S., Degenhardt, L., Hoban, B. & Gisev, N. A synthesis of oral morphine equivalents (OME) for opioid utilisation studies. Pharmacoepidemiol. Drug Saf. 25, 733–737 (2016).

17. D’Souza, G. et al. Pharmacological strategies for decreasing opioid therapy and management of side effects from chronic use. Children (Basel) 5, 163 (2018).

18. Abie Horowitz, M., Framer, A., Strang, J. & Taylor, D. Tapering and withdrawing opioids: guidance informed by fundamental principles to minimise withdrawal symptoms. Ther. Adv. Psychopharmacol. 15, 20451253251371504 (2025).

19. Bruera, E. & Paice, J. A. Cancer pain management: safe and effective use of opioids. Am. Soc. Clin. Oncol. Educ. Book e593–9 (2015).

20. https://www.hca.wa.gov/assets/billers-and-providers/pharmacy-opioid-quick-guide.pdf.

21. https://www.fda.gov/media/147152/download.

22. Goodell, A. J., Chu, S. N., Rouholiman, D. & Chu, L. F. Large language model agents can use tools to perform clinical calculations. NPJ Digit. Med. 8, 163 (2025).

23. DiPrete, B. L. et al. Association of opioid dose reduction with opioid overdose and opioid use disorder among patients receiving high-dose, long-term opioid therapy in North Carolina. *JAMA Netw*. Open 5, e229191 (2022).

24. Webster, L. R. & Fine, P. G. Review and critique of opioid rotation practices and associated risks of toxicity. Pain Med. 13, 562–570 (2012).

25. Telenti, A. et al. Large language models for science and medicine. Eur. J. Clin. Invest. 54, e14183 (2024).

26. Ganjavi, C. et al. ChatGPT and large language models (LLMs) awareness and use. A prospective cross-sectional survey of U.S. medical students. PLOS Digit. Health 3, e0000596 (2024).

27. Xu, A. Y. et al. A pilot study of medical student opinions on large language models. Cureus 16, e71946 (2024).

28. Savage, T., Nayak, A., Gallo, R., Rangan, E. & Chen, J. H. Diagnostic reasoning prompts reveal the potential for large language model interpretability in medicine. NPJ Digit. Med. 7, 20 (2024).

29. Chen, S. et al. When helpfulness backfires: LLMs and the risk of false medical information due to sycophantic behavior. NPJ Digit. Med. 8, 605 (2025).

30. Park, S. H. et al. Minimum reporting items for CLear Evaluation of Accuracy Reports of Large Language Models in healthcare (MI-CLEAR-LLM): 2025 updates. Korean J. Radiol. 26, 1123–1132 (2025).

